# Antibiotic resistance in clinical isolates of *Escherichia coli* non-producer of Extended Spectrum Betalactamases (ESBL), obtained from urine cultures in patients with urinary tract infection

**DOI:** 10.1101/2020.08.15.20173146

**Authors:** Jorge Angel Almeida Villegas, Mariana Islas Rodriguez, Iris Mellolzy Estrada, Miriam Deyanira Rodriguez, María Carolina Erazo Muñoz, Silvia Patricia Rodriguez

## Abstract

**Objective:** To identify strains of *Escherichia coli* that do not produce Extended Spectrum Betalactamases (ESBL) in urine cultures and to evaluate the patterns of resistance to certain antibiotics used in clinical practice as treatments of choice in the Toluca Valley of Mexico.

**Method:** 155 urine samples were obtained from people in the Toluca Valley, Mexico under necessary hygiene conditions. 100ml used to identify the pathogen and its resistance patterns were collected from each sample, using an automated Walkaway method.

**Result:** 19 strains of non-ESBL-producing *Escherichia coli* were obtained, of which 68.42% showed resistance to ampicillin and tetracycline, 47.36% resistance to trimethoprim with sulfamethoxazole, 26.31% for levofloxacin, ciprofloxacin, ceftriaxone, and resistance of 21 % with ampicillin and sulbactam. 100% sensitivity for amikacin, tigecycline and carbapenems.

**Conclusion:** Antibiotic resistance represents a major health problem, as the present study shows, ampicillin is a drug of choice for urinary tract infection produced *by Eschericha coli*, but in the Toluca Valley the resistance to this antibiotic is 68.42% In non-ESBL producing strains, high resistance patterns are also shown for quinolones, tetracycline and trimethoprim with sulfamethoxazole.

## Introduction

The urethra is a portal for the exit of urine, but also allows the entry of microbes, including pathogens, into the urinary tract.^1^ The phrase “urinary tract infection” (UTI) is a general term that refers to an infection, usually bacterial in etiology, anywhere along the urinary tract from the urethral meatus to the perinephric fascia. Structures in this pathway include the urethra, bladder, ureters, and the renal pelvis and parenchyma. Associated structures that also become infected and that may serve as foci of recurrent UTI are the prostate, epididymis, and perinephric fascia. Specific types of urinary tract infection include urethritis, an infection limited to the urethra; cystitis, an infection of the bladder; and pyelonephritis, a more extensive infection involving the upper urinary tract structures.^2^

Symptomatic urinary tract infection in a healthy is a complex event. It is initi-ated when potential urinary pathogens from the bowel, s colonize the periurethral mucosa and ascend through the urethra to the bladder and in some cases through the ureter to the kidney.^3^

Urinary tract infection (UTI) is one of the most commonproblems seen by primary care physician.^4^ The microbial etiology of urinary infections has for several decades been regarded as well established. *Escherichia coli* remains the predominant uropathogen isolated. The etiology of UTI is also affected by underlying host factors that complicate UTI, such as age, diabetes, spinal cord injury, or catheterization. Consequently, complicated UTI has a more diverse etiology tan uncomplicated UTI, and organisms that rarely cause disease in healthy patients can causes ignificant disease in hosts with anatomic, metabolic,or immunologic underlying disease.^5^ Urinary tract infection is mainly caused by gram-negative organisms that include *E. coli, Klebsiella, Proteus*, and *Pseudomonas*, and gram-positive bacteria, group B *Streptococcus* and *Staphylococcus* species.^6^

Antibiograms are usually performed when empirical therapy fails. Several studies have shown a high prevalence of multidrug resistance in hospital and community-based *E. coli* uropathogens.^7^ Surveillance studies provide information of the causativeagents of UTIs and their antimicrobial resistancepatterns which may aid clinicians in choosing theappropriate antimicrobial empirical treatment.^8^

Ampicillin has been frequently used for urinary tract infection as an alternative drug owing to its activity against gram-negative microorganisms. Nonetheless, as acquired resistance to betalactam class antibiotics was reported, cephalosporins stable to betalactamase were developed, and thus the choice of antibiotics for urinary tract infection was widened.^9^

The introduction of antimicrobial therapy has contributed significantly to the management of UTIs. However the main problem with current antibiotic therapies is the rapid emergence of antimicrobial resistance in hospitals and the community.^10^ Currently several bacterial strains are resistant to practically all known antibiotics. Such as carbapenems, cephalosporins, macrolides, and penicillins.^11^

## Method

In the present study, 155 urine cultures were performed in patients with sintom clinical to urinary tract infection, without exclusion criteria such as gender or age.

Of the 155 urine cultures performed, the distribution was; 107 people of the female gender representing 69% of the study population and 48 men representing 31%. The age ranges in the lower limit is a 1-year-old and for the higher limit is a 91-year-old patient, with an average of 25 to 30 years.

For the study, 100 ml of the first urine of the day were collected with the necessary hygiene conditions for the collection of the specimen.

An aliquot of urine was taken with a calibrated handle, which was plated on chromogenic agar for cross streak and on agar agar for dispersion. The plates were incubated 24 hours at 37 ° C.

Colony count of blood agar medium was performed. An isolated colony was taken from the chromogenic agar and diluted in diluent medium for plate reseeding for automated equipment.

The identification of the etiological agent as well as the sensitivity tests were performed using an automated method using the walkaway SI 96 from beckman coulter equipment.

### Result

Among the 155 urine cultures performed, 80 were positive for microbial development and 75 showed no growth.

The 51.62% that represents positive cases has a distribution that 47.5% represents bacteria other than *E coli*, 8.75% corresponds to yeasts and 43.75% represents *E coli*, which is equivalent to 35 cultures. Of the 35 *Escherichia coli* isolates, 19 strains are not ESBL producing, which corresponds to 54.28% of *E coli* isolates.

Table 1 shows the patterns of resistance to the different antibiotics used in the clinical isolates of *E. coli*.

**Table 1.** The following table shows the antibiotics used, as well as the designations of resistant, intermediate sensitivity and sensitive. Placing the 19 strains isolated in each space according to resistance pattern

| Antibiotic | Sensitivity patterns |  |  |
| --- | --- | --- | --- |
|  | Resistant | Intermediate Sensitivity | Sensitive |
| Amikacin | 0 | 0 | 19 |
| Amoxicillin /Clavulanic Acid | 2 | 1 | 16 |
| Ampicillin | 13 | 1 | 5 |
| Ampicillin /Sulbactam | 4 | 4 | 11 |
| Cefazolin | 5 | 0 | 14 |
| Cefepime | 1 | 3 | 15 |
| Cefotaxime | 2 | 1 | 16 |
| Ceftazidime | 3 | 1 | 15 |
| Ceftriaxone | 5 | 0 | 14 |
| Cefuroxime | 5 | 0 | 14 |
| Ciprofloxacin | 5 | 0 | 14 |
| Ertapenem | 0 | 0 | 19 |
| Gentamicin | 3 | 1 | 16 |
| Imipenem | 0 | 0 | 19 |
| Levofloxacin | 5 | 2 | 12 |
| Meropenem | 0 | 0 | 19 |
| Piperacillin/ Tazobactam | 0 | 0 | 19 |
| Tetracycline | 13 | 0 | 6 |
| Tigecycline | 0 | 0 | 19 |
| Tobramycin | 4 | 2 | 13 |
| Trimethoprim/Sulfamethoxazole | 9 | 0 | 10 |

The isolated strains showed different patterns of resistance, in which the greatest resistance was found in betalactamic ampicillin with a pattern of 68.42%. When ampicillin is used with sulbactam the resistance is only 21.05%, this pattern does not decrease significantly.

Tetracycline is matched with ampicillin with a resistance of 68.42% and second with a high number of resistance is trimethoprim with sultamethoxazole with 47.36%.

In the case of quinolones such as ciprofloxacin and levofloxacin, as well as for cephalosporins such as cefuroxime, ceftriaxone and cefzolin, the resistance pattern is 26.31%

Antibiotics such as aminoglycoside amikacin, carbapenems imipenem, ertapenem, and meropenem, as well as piperacillin with tazobactam and tigecycline have a sensitivity pattern of 100%.

Graph 1 shows the resistance patterns as well as the number of strains in each pattern.

**Graph 1.**
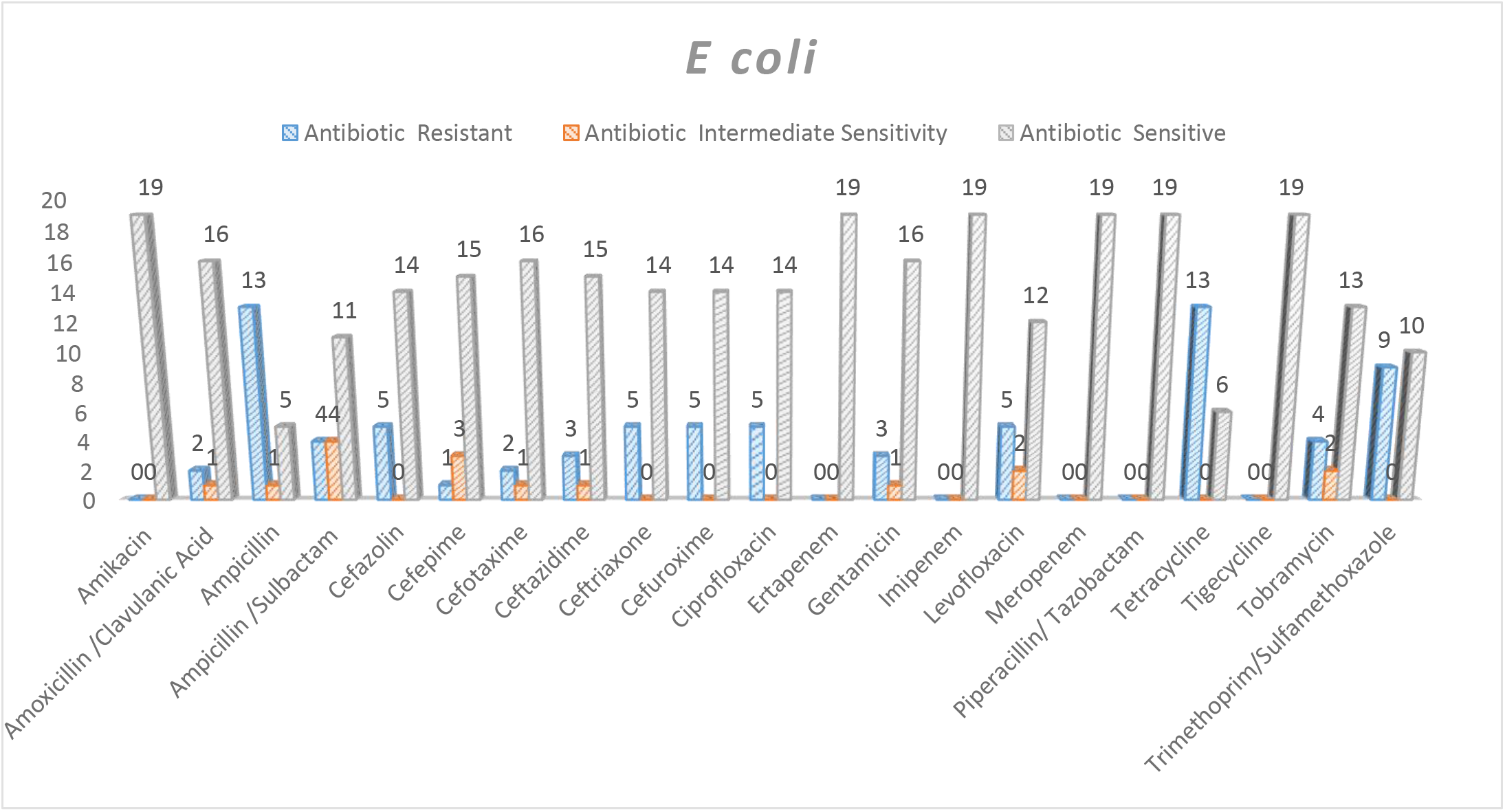
Shows the patterns for the different antibiotics and the number of strains found in the different patterns under study. In blue for resistant strains, orange for intermediate sensitivity and gray for sensitive strains.

## Discusion

Enterobacteriaceae such as *Escherichia coli* are considered betalactamase, extended spectrum betalactamase, and carbapenemase-producing bacteria responsible for the inactivation of the betalactam ring of betalactam antibiotics. This explains the high percentage of resistance to antibiotics such as ampicillin with 68.42%, the same percentage that is reduced in ampicillin with subactam, this due to the presence of sulbactam, a betalactam inhibitor, which confers a greater therapeutic effect of the antibiotic. However, 68.42% ampicillin and 21.05% ampicillin with sulbactam suggest the presence of betalactamases. In the case of cephalosporins, ceftriaxone, cefuroxime and cefazolin that show a resistance of 26.31%, betalactamases, mainly of extended spectrum betalactamases, can also be suspected.

The presence of ESBL also allows the acquisition of resistance mechanisms to other antibiotics, such as the case of quinolones, where levofloxacin and ciprofloxacin have the same percentage of resistance as second and third generation cephalosporins, with 26.31% and a high pattern. Tetracycline resistance with 68.42%.

Both betalactamases and extended spectrum betalactamases have no effect on carbapenems, so the sensitivity pattern to ethos was 100%.

## Conclusion

Antibiotic resistance has increased markedly, making it difficult to treat infections such as UTIs, ampicillin is one of the drugs of choice for this condition, however, the study shows that in the Toluca Valley, Mexico, ESLE non-ESBL-producing strains of *Escherichia coli* show high resistance with 68.42% of isolates. As well as, marked resistance to trimethoprim with sulfamethoxazole and tetracycline.

Medications such as amoxicillin with clavulanic acid, cephalosporins, amikacin, gentamicin, and carbapenems, still represent a suitable treatment for UTI caused by *Escherichia coli* not producing ESBL.

The study suggests a specific determination of the betalactamases present in the strains, in order to clarify the exact mechanism of resistance.

## Data Availability

Readers will be able to access the data here present, citing and giving credit to the authors of this research work.

## Interest Conflict

The authors declare that they have no conflict of interest.

## Acknowledgment

We thank all the institutions that allowed us to carry out this research work.

## Financing

No funding was received to carry out this work.

## Notes

### Competing Interest Statement

The authors have declared no competing interest.

### Funding Statement

The authors declare that they have no conflict of interest.
No funding was received to carry out this work.

### Author Declarations

The Microtec Laboratories Ethics Committee has indicated that the research project does not need approval.

